# Evidence for a pervasive autobiographical memory impairment in Logopenic Progressive Aphasia: clinical and neural correlates

**DOI:** 10.1101/2020.06.14.20131383

**Authors:** Siddharth Ramanan, David Foxe, Hashim El-Omar, Rebekah M. Ahmed, John R. Hodges, Olivier Piguet, Muireann Irish

## Abstract

Logopenic Progressive Aphasia is a rare language disorder characterised by repetition and naming difficulties, reflecting the progressive degeneration of left-lateralized peri-sylvian temporal and inferior parietal regions. Mounting evidence suggests that cognitive impairments in this syndrome extend beyond the language domain to include episodic encoding and retrieval disturbances. To date, it remains unknown whether autobiographical memories from across the lifespan are also subject to decline, yet this information is critical to arrive at a comprehensive understanding of the Logopenic syndrome. The objective of this study was to provide the first in depth examination of autobiographical memory function in Logopenic Progressive Aphasia using the Autobiographical Interview, a validated semi-structured interview which assesses recollection of the past under free and probed recall conditions. Autobiographical memory performance in 10 well-characterised Logopenic Progressive Aphasia patients was contrasted with that of 18 typical amnestic Alzheimer’s disease and 16 healthy Control participants. Relative to Controls, Logopenic Progressive Aphasia cases showed marked impairment in the free recall of episodic details, scoring comparably to disease-matched cases of Alzheimer’s disease. This impairment was evident across all time periods and persisted even when formal structured probing was provided. Importantly, controlling for overall level of language disruption failed to ameliorate the autobiographical memory impairment in the Logopenic Progressive Aphasia group, suggesting a genuine amnesia spanning recent and remote memories. Whole-brain voxel-based morphometry analyses revealed that total episodic information retrieved in Logopenic Progressive Aphasia was associated with decreased grey matter intensity predominantly in a bilateral posterior parietal network. Taken together, our findings reveal for the first time the presence of marked remote and recent autobiographical memory impairments in Logopenic Progressive Aphasia, that cannot be explained solely due to their language difficulties or disease staging. Our findings hold important clinical implications for the accurate characterization of Logopenic Progressive Aphasia, and suggest that episodic memory difficulties should be considered as one of the core clinical features of this syndrome.

## INTRODUCTION

Logopenic Progressive Aphasia (LPA) is a rare neurodegenerative clinical syndrome characterised by marked reductions in spontaneous speech, in the context of phonological errors, word-finding, and sentence repetition difficulties (Gorno-Tempini *et al*., 2008; Gorno-Tempini *et al*., 2011). This pattern of language dysfunction is largely held to reflect the disruption of phonological working memory and lexical retrieval functions posited to support repetition, and spontaneous speech and naming, respectively (Henry and Gorno-Tempini, 2010; Leyton *et al*., 2012). Grammatical processing and semantic comprehension abilities in LPA, by contrast, are relatively preserved until later stages of the disease (Gorno-Tempini *et al*., 2004; Gorno-Tempini *et al*., 2008). Neuroanatomically, the epicentre of atrophy in LPA resides in the inferior parietal cortex (IPL), superior/middle temporal gyri and adjacent perisylvian cortices (Gorno-Tempini *et al*., 2008; Rohrer *et al*., 2010; Leyton *et al*., 2012; Teichmann *et al*., 2013). Evidence for medial temporal and hippocampal dysfunction, by contrast, appears mixed, with some studies reporting early degradation of the hippocampus (Gorno-Tempini *et al*., 2004; Rohrer *et al*., 2013; Phillips *et al*., 2019), while others note its relative preservation until later in the disease course (Teichmann *et al*., 2013; Win *et al*., 2017).

Although characterised primarily as a disorder of language, mounting evidence points to multidimensional non-linguistic cognitive impairments in LPA, including spatial working memory, visuospatial and executive functions (Foxe *et al*., 2013; Butts *et al*., 2015; Watson *et al*., 2018; Kamath *et al*., 2019; Mendez *et al*., 2019; Ramanan *et al*., 2019a). Of particular interest is recent work demonstrating the presence of marked episodic memory difficulties in LPA on neuropsychological tests of verbal (Piguet *et al*., 2015; Win *et al*., 2017; Eikelboom *et al*., 2018) and nonverbal (Piguet *et al*., 2015; Ramanan *et al*., 2016) episodic delayed recall. These objective impairments, in turn, are corroborated by subjective patient and carer reports of everyday memory difficulties (Magnin *et al*., 2013; Ramanan *et al*., 2016), suggesting that episodic memory disturbances may be an under-appreciated feature of this disorder.

An important issue to address is whether the verbal demands of episodic memory tasks give rise to a spurious memory impairment that is best accounted by dysfunctional lexical retrieval (Win *et al*., 2017), rather than a core memory impairment, *per se*. The finding of pronounced impairments on non-verbal assays of episodic memory in LPA, however, suggests the presence of a genuine retrograde amnesia in this group (Ramanan *et al*., 2016; Santos-Santos *et al*., 2018; Mendez *et al*., 2019). We recently demonstrated that verbal and nonverbal episodic retrieval deficits in LPA are attributable, in part, to dysfunction of two key memory processing regions, namely the angular gyrus in the left IPL and the left posterior hippocampus, as well as white matter tracts connecting these regions (Ramanan *et al*., 2020). Independent studies, moreover, indicate that hypometabolism of parietal and IPL regions, as opposed to temporal lobe-dominant profiles, are strongly associated with verbal and visual memory difficulties in LPA (Krishnan *et al*., 2017). Collectively, these findings point to the presence of incipient episodic amnesia in LPA, most likely reflecting the degeneration of posterior parietal brain regions (Sestieri *et al*., 2017).

What remains unclear is whether memory difficulties in LPA can be attributed to post-disease onset factors, such as poor encoding. Examining the status of memories encoded *prior* to disease onset provides an important window into the nature and severity of memory impairment in neurological disorders. Here, we focus on autobiographical memory; the recollection of personally-relevant experiences replete with rich sensory-perceptual, emotional, and semantic information (Conway, 2001; Irish and Piguet, 2013; D’Argembeau, 2020). Autobiographical memory is a particularly fruitful aspect of memory to explore given the availability of well-validated interview techniques that have been reliably used in neurodegenerative populations (see, for example McKinnon *et al*., 2006; McKinnon *et al*., 2008; Irish *et al*., 2011a; Hsieh *et al*., 2016; Ahmed *et al*., 2018; Irish *et al*., 2018; Carmichael *et al*., 2019). Moreover, the neural substrates of autobiographical memory retrieval are well-defined, with a large corpus of functional neuroimaging studies demonstrating the involvement of a ‘core memory network’, anchored on the hippocampus, that also includes IPL, medial parietal and frontotemporal regions (Svoboda *et al*., 2006; Chen *et al*., 2017; Boccia *et al*., 2019). Lesion studies consistently show that damage to any of the nodes of this core memory network negatively impacts the episodic and autobiographical recollective endeavour (Levine, 2004; Berryhill, 2012; Irish *et al*., 2012; McCormick *et al*., 2018; Ramanan *et al*., 2018). Of particular interest for the current study, patients with focal parietal lesions (centred on the IPL) display marked difficulties in the spontaneous retrieval of autobiographical memories across the entire lifespan, producing event narratives that are divested of sensory-perceptual and contextual information (Berryhill *et al*., 2007; Davidson *et al*., 2008). Neurostimulation studies further demonstrate that functional inhibition of the IPL negatively impacts the retrieval of episodic information from autobiographical memory (Thakral *et al*., 2017; Bonnici *et al*., 2018). As such, there is growing consensus that parietal lobe dysfunction can result in stark amnesia (Ramanan *et al*., 2018).

While autobiographical memory disturbances are well-documented in Alzheimer’s disease (AD) using a variety of techniques (e.g., Irish *et al*., 2011b; Barnabe *et al*., 2012), no study has assessed the capacity for autobiographical retrieval in LPA. This gap in the literature is surprising given unequivocal evidence for marked disturbances in both episodic learning (Casaletto *et al*., 2017) and retrieval (Ramanan *et al*., 2016; Win *et al*., 2017; Eikelboom *et al*., 2018; Ramanan *et al*., 2020) in this syndrome. The objective of the current study was to provide a fine-grained examination of autobiographical memory retrieval in LPA using a widely-used and validated assessment tool – the Autobiographical Interview (Levine *et al*., 2002). This measure assesses event retrieval from recent and remote life epochs under conditions of minimal retrieval support (free recall) versus high retrieval support (structured probing) and permits the content of autobiographical narratives to be parsed into discrete contextual detail profiles. We hypothesised that autobiographical memory impairments would be present in LPA and of the same magnitude as that observed in disease-matched cases of AD, extending across the entire lifespan to affect recent and remote experiences comparably. Importantly, we predicted that autobiographical memory impairment in LPA would not be primarily accounted for by the characteristic language disturbances of this disorder. Moreover, the provision of structured probing would not be sufficient to ameliorate memory deficits in LPA, suggesting a genuine episodic memory impairment. Finally, using whole-brain voxel-based morphometry analyses, we sought to determine the neural substrates of autobiographical memory impairments in LPA. In this regard, we predicted that degeneration of the left IPL would emerge as a core determinant of autobiographical memory impairment in LPA, underscoring the role of this region in mnemonic functions (Ramanan *et al*., 2018; Rugg and King, 2018).

## MATERIALS AND METHODS

### Participants

A total of 44 participants were recruited through FRONTIER, the frontotemporal dementia research group at the Brain and Mind Centre, The University of Sydney, Australia. Ten patients with a clinical diagnosis of LPA, characterised by naming, sentence and single word repetition and word finding difficulties, in the absence of motor speech deficits or semantic knowledge impairment, were included (Gorno-Tempini *et al*., 2011). Data from carer-reported history and clinical examination of language performance were used to corroborate the diagnosis of LPA (see Supplementary Table 1). As a disease-control group, we included 18 patients with a clinical diagnosis of probable AD with a predominantly amnestic presentation (McKhann *et al*., 2011). Atypical variants of AD such as dysexecutive AD or Posterior Cortical Atrophy were excluded.

Diagnoses of LPA and AD were established by consensus among a multidisciplinary team comprising senior neurologists (R.A., J.R.H.), a clinical neuropsychologist, and an occupational therapist based on comprehensive clinical and neuropsychological assessment and structural neuroimaging evidence in line with current diagnostic criteria for both syndromes. Disease severity was indexed using the clinician-rated Clinical Dementia Rating – Frontotemporal Lobar Degeneration – Sum of Boxes (CDR-FTLD SoB: Knopman *et al*., 2008) measure, which provides a global impression of changes in cognition, behaviour, and activities of daily living. Carers completed the Cambridge Behavioural Inventory – Revised (CBI-R; Wear *et al*., 2008) to assess behavioural changes in patients.

Sixteen healthy older control participants were selected through the FRONTIER research volunteer panel. All controls scored 88 or above on the Addenbrooke’s Cognitive Examination – Revised (ACE-R; Mioshi *et al*., 2006) or its updated counterpart, the ACE-III (Hsieh *et al*., 2013) – both of which are short and comparable assessments of global cognitive functions measuring attention and orientation, memory, language, verbal fluency, and visuospatial processing abilities (see So *et al*., 2018). Healthy controls scored 0 on the CDR-FTLD SoB measure. Participants with a history of significant head injury, cerebrovascular disease, substance abuse, other primary psychiatric, mood, or neurological disorders, or limited English proficiency were excluded.

All participants or their Person Responsible provided written informed consent in accordance with the Declaration of Helsinki. The study was approved by The University of New South Wales and South Eastern Sydney Local Health District ethics committees.

### General and targeted neuropsychological assessment

All participants underwent comprehensive neuropsychological assessment of language, memory, and executive function. Due to a change in testing protocol, five LPA patients underwent the ACE-III and for uniformity, their scores were converted to ACE-R scores, using a validated conversion algorithm specific to the LPA syndrome (see So *et al*., 2018). The ACE-R total score (max score = 100; Mioshi *et al*., 2006; Hsieh *et al*., 2013) was used as an index of overall cognitive performance, with total scores from its Memory and Language subdomains (max score for both = 26) used as indices of memory and language performance, respectively. In addition to general neuropsychological screening measures, targeted assessments of cognitive performance were used to assess language (including sentence repetition, single word repetition, naming, semantic association), attention and executive, visuospatial, and episodic memory performance. For brevity, the administration and scoring details for these assessments are presented in Supplementary Material.

### Assessment of autobiographical memory

Autobiographical memory was assessed using a shortened version of the Autobiographical Interview (Levine *et al*., 2002) that has been employed across a range of neurodegenerative dementia cohorts (e.g. McKinnon *et al*., 2006; McKinnon *et al*., 2008; Irish *et al*., 2011a; Irish *et al*., 2018; Seixas Lima *et al*., 2019). It should be noted that autobiographical memory data from the AD group have been previously presented elsewhere (see e.g., Irish *et al*., 2011a) and are included here purely as a disease comparison to the LPA group.

Autobiographical memory performance was examined across four time periods: Teenage (11-17 years), Early Adulthood (18-34 years), Middle Adulthood (35-55 years), and Recent past (within the last year). Briefly, all participants were instructed to provide a detailed verbal description of a personally experienced event that occurred at a specific time and place from each time period. In line with the original administration protocol, the level of retrieval structure was manipulated across three conditions: Free Recall, General Probe, and Specific Probe. The Specific Probe condition was administered after all events had been retrieved via Free Recall and General Probe conditions to avoid contaminating free recall of subsequent memories (Levine *et al*., 2002). Interviews for all participants were digitally recorded for subsequent transcription and scoring.

### Scoring of autobiographical memories

Autobiographical memories were scored according to the standard Autobiographical Interview protocol (Levine *et al*., 2002). First, retrieved information was segmented into informational bits or details and categorised as ‘internal’ or ‘external’. Internal details related to the main event of interest, and formed the key metric of interest here reflecting episodic recollection. These details were further assigned to one of five separate contextual detail subcategories (Event, Time, Place, Perceptual, and Thought/Emotion). External details largely reflect details tangential to the main event, repetitions, metacognitive statements, or semantic facts (but see also Strikwerda-Brown *et al*., 2018). Details were summed to form composite internal and external details scores separately for each condition (Free Recall, General Probe, Specific Probe). As the General Probe condition elicits minimal appreciable effects on recall performance, output from the Free Recall and General Probe conditions was combined (hereon referred to as Free Recall; Levine *et al*., 2002). A Total Recall score was also computed (i.e., sum of Free Recall, General Recall, and Specific Probe conditions) reflecting the total amount of autobiographical information retrieved, following provision of structured probes. Preliminary analyses of missing data revealed one missing data point (comprising ∼0.5% of the total Autobiographical Interview data) for a single LPA patient for the Middle Adulthood period. This score was imputed with the median score of the Middle Adulthood epoch from the LPA group (see Supplementary Methods). In the case of word-finding difficulties or circumlocuitous speech, participants were credited with a point for each meaningful detailed item. This was to ensure that LPA patients were not unfairly penalized for semantic word retrieval difficulties (see also Statistical analyses section on controlling for overall language dysfunction). All interviews were scored by an experienced rater, blind to group and study hypotheses (D.F.).

### Statistical analyses

Behavioural data were analysed using RStudio v3.3.0 (R Core Team, 2016). First, Shapiro-Wilk tests and box-and-whisker plots were used to assess whether data were normally distributed. Accordingly, chi-square (χ^2^) tests (examining group differences in sex), *t*-tests (investigating group differences in age of disease onset and disease severity), analyses of variance (ANOVA; examining group differences in age, education, and neuropsychological measures) were employed. Analyses of Covariance (ANCOVA) were used to explore group differences on the Autobiographical Interview, controlling for overall language performance on the ACE-R Language subscale, to account for primary language difficulties in the LPA group. The main outcome measures on the Autobiographical Interview included: (i) total retrieval of internal details in Free Recall and Total Recall conditions, summed across time periods, (ii) the ratio of internal details to overall information (i.e., sum of internal and external details) to understand the proportion of episodic information produced within the overall autobiographical narrative, and (iii) profiles of contextual information retrieved across internal detail subcategories (i.e., Event, Time, Place, Perceptual, and Thought/Emotion details).

Partial eta-squared values 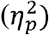 accompanied all ANOVAs and ANCOVAs as measures of corresponding effect sizes. *Post-hoc* comparisons between LPA, AD, and Control groups were corrected using Sidak corrections for small sample sizes. Finally, Pearson’s correlations (*r* values) were employed to examine associations between internal details produced during Free Recall and Total Recall conditions, disease severity, carer-rated memory difficulties, and neuropsychological test performance in LPA and AD groups separately. For correlations, all *p*-values were corrected for multiple comparisons using the ‘false discovery rate’ approach (Benjamini and Hochberg, 1995).

### Epoch-specific analyses

Repeated-measures ANCOVAs, covarying for overall language performance, were employed to explore main effects of group and epoch for internal details, as well as relevant interactions (Levine *et al*., 2002; McKinnon *et al*., 2008). Separate repeated-measures ANCOVAs were run for Free Recall and Total Recall conditions as the level of Probed detail hinges upon the number of Free recall details produced (i.e., the two scores are not independent). The resulting *p*-values were subject to Greenhouse-Geisser corrections for sphericity. Generalised eta-square 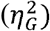 effect size metrics accompanied all repeated-measures ANCOVA results (Bakeman, 2005).

### Image acquisition

Forty-one participants (10 LPA, 18 AD and 13 Controls) underwent whole-brain T_1_-weighted structural brain magnetic resonance imaging (MRI) using a 3T Philips scanner with standard quadrature head coil (eight channels). The 3D T_1_-weighted images were acquired using the following sequences: coronal acquisition, matrix 256 × 256 mm, 200 slices, 1mm isotropic voxel resolution, echo time/repetition = 2.6/5.8 msec, flip angle α=8°.

### Voxel-based morphometry

Whole brain voxel-based morphometry (VBM) analyses were employed to investigate voxel-by-voxel changes in grey matter intensity between groups, using FSL (FMRIB Software Library: https://fsl.fmrib.ox.ac.uk/fsl/fslwiki). Briefly, a standard pre-processing pipeline was employed involving brain extraction (Smith, 2002), tissue segmentation (Zhang *et al*., 2001), and non-linear registration (Andersson *et al*., 2007a, b) to align brain-extracted images to the Montreal Neurological Institute (MNI) standard space. Full details of pre-processing steps are provided in Supplementary Methods.

To investigate whole-brain voxel-wise grey matter intensity differences between LPA, AD, and Control groups, independent *t*-tests were run with age included as a nuisance variable (see Supplementary Methods). Clusters were extracted using the Threshold-Free Cluster Enhancement method using a threshold of *p* < .005 corrected for Family-Wise Error with a cluster threshold of 100 contiguous voxels.

Correlation analyses were run to determine the relationship between whole-brain grey matter intensity and the total number of internal details (summed across all time periods) produced on the Autobiographical Interview. Separate general linear models were run for Free Recall and Total Recall conditions, using a covariate-only statistical model with a positive *t*-contrast. Correlation analyses were performed for each patient group combined with the Control group with age and overall language performance (measured using ACE-R Language subscale score) included as nuisance variables. To ensure that correlations were not driven by a bimodal distribution, the mode and mean scores for total internal details in Free Recall and Total Recall conditions in the combined LPA-Control group were extracted and visually inspected via histogram distributions (see also Bertoux *et al*., 2014). For both contrasts, a single mode that was different from the combined group’s mean underlay the distribution (Supplementary Table 2), indicating a unimodal distribution for internal details. Anatomical locations of statistical significance were overlaid on the MNI standard brain with maximum coordinates provided in MNI stereotaxic space. Clusters were extracted using a strict threshold of *p* < .001 uncorrected for multiple comparisons with a cluster extent threshold of 50 contiguous voxels to capture changes in subcortical striatal and medial temporal regions whilst mitigating Type I and Type II errors (Lieberman and Cunningham, 2009).

### Data availability

The ethical requirement to ensure patient confidentiality precludes public archiving of our data. Researchers who would like to access the raw data should contact the corresponding author who will liaise with the ethics committee that approved the study, and accordingly, as much data that is required to reproduce the results will be released to the individual researcher.

## RESULTS

### Demographic and clinical variables

Demographic and clinical characteristics for all participants are presented in Table 1. Participant groups were comparable in terms of age and education (both *p* values > .1), however, differences in sex distribution were evident with more males in the AD group (77%) compared to more females (76%) in the Control group (*p* = .01). These sex differences did not significantly impact autobiographical retrieval performance on either Free or Total Recall conditions (Supplementary Table 3) and are, therefore, not considered further. Importantly, both patient groups were comparable for age at disease onset, clinician-indexed disease severity (CDR-FTLD SoB), and carer-reported changes in behaviour and memory (CBI-R: all *p* values > .1). LPA and AD groups scored significantly lower on the ACE-R total than Controls (both *p* values < .001), with no significant differences between the patient groups (*p* > .1). These data indicate comparable levels of overall cognitive dysfunction and disease staging in the LPA and AD groups.

**Table 1.**
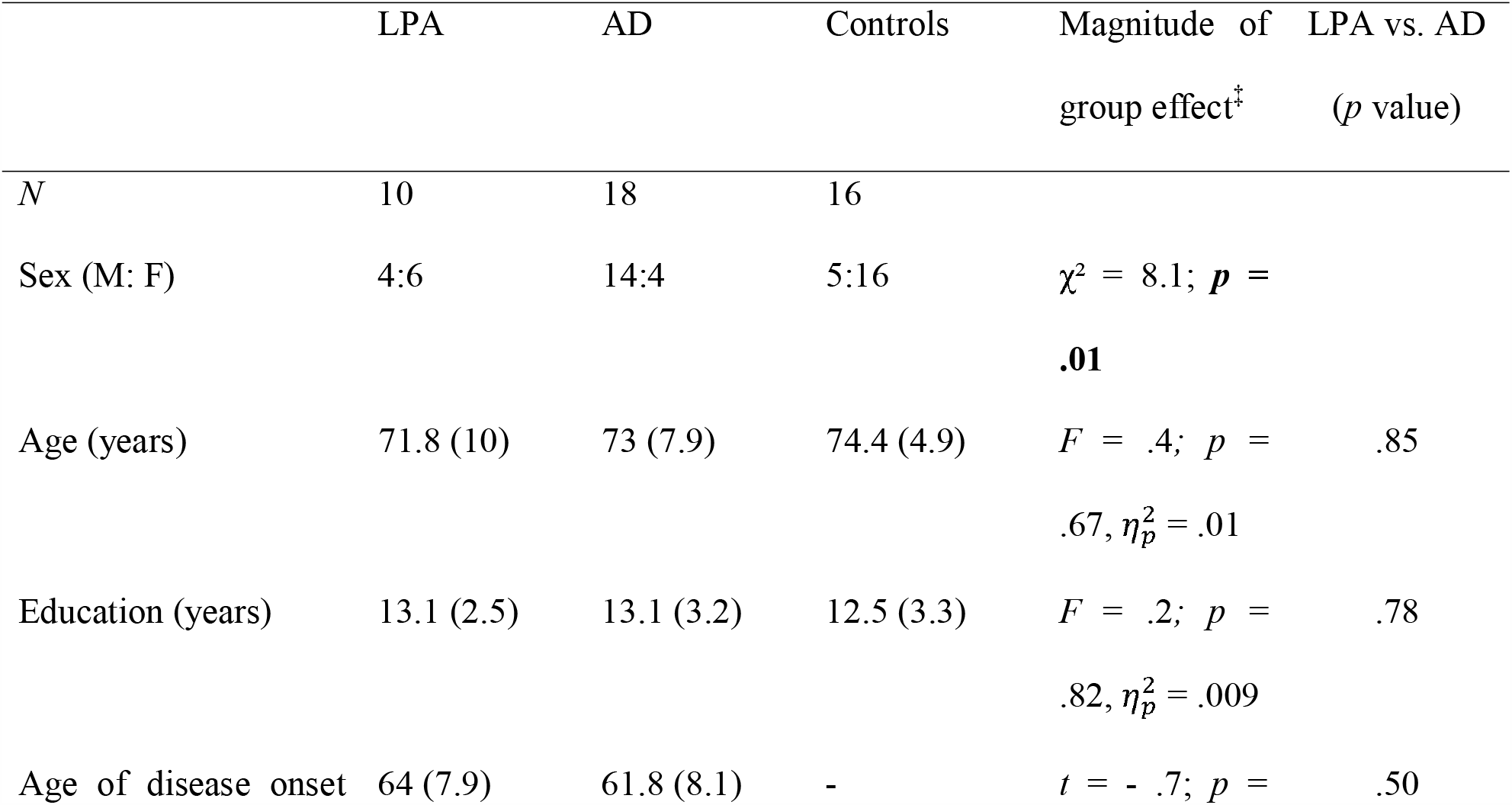
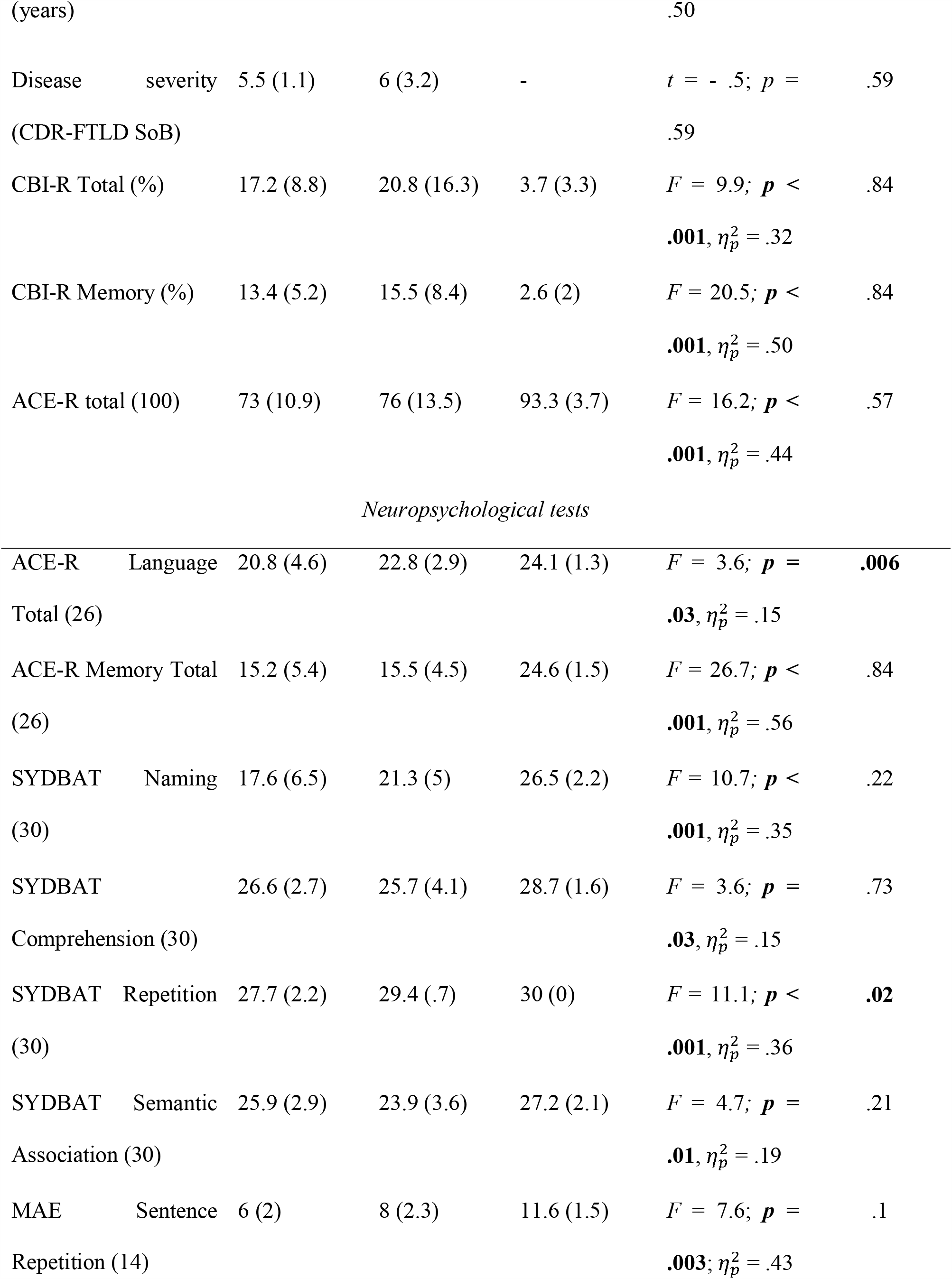
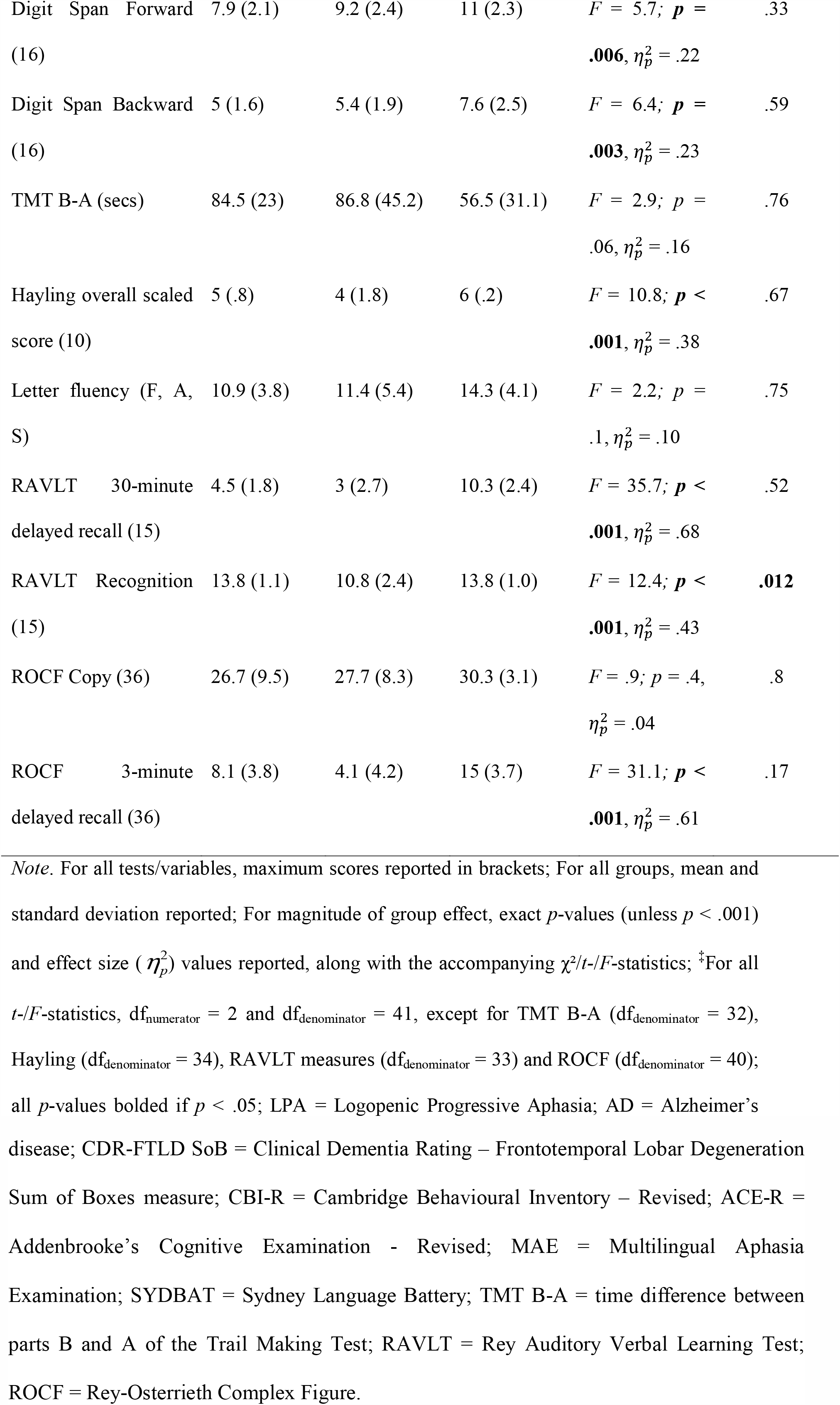
Demographic, clinical, and general neuropsychological assessment performance for all groups

### Neuropsychological test performance

Neuropsychological testing revealed characteristic profiles of cognitive impairment in LPA and AD (Table 1). Relative to Controls, the LPA group demonstrated significant verbal and nonverbal episodic memory and language impairments, as well as the canonical profile of language difficulties spanning confrontational naming, comprehension, single word and sentence repetition, auditory attention and working memory (all *p* values ≤ .03). The AD group displayed significant episodic memory dysfunction in the context of semantic processing impairments, and difficulties in working memory and executive function (all *p* values < .05), in comparison with Controls. Direct contrasts between patient groups revealed disproportionate impairments in language, specifically single-word repetition and overall language performance on the ACE-R Language subscale in LPA relative to AD, while AD patients displayed greater impairments in verbal recognition memory (measured using the Rey Auditory Verbal Learning Test Recognition component) relative to LPA (all *p* values < .05; Table 1).

### Autobiographical memory performance

#### Overall Free Recall

Table 2 and Figure 1A display the group-level performance for Free Recall of internal details summed across time periods. An ANCOVA controlling for language function revealed a significant main effect of group [*F* (2, 40) = 14.2; *p* < .001; 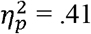], with both LPA and AD groups retrieving fewer internal details relative to Controls (both *p* values < .005; Figure 1A), and no significant difference between the patient groups (*p* > .1). The ratio of internal-to-total detail production, controlling for language dysfunction, further revealed an overall main effect of group [*F* (2, 40) = 17.6; 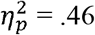], with LPA and AD patients producing a lower proportion of internal (episodic) details within the overall autobiographical narrative compared to Controls (both *p* values < .001) (Supplementary Table 4). The proportion of internal details provided during Free Recall did not differ between the patient groups (*p* = .79) (Supplementary Table 4). Looking more closely at the type of internal details produced during Free Recall, the LPA group produced significantly fewer Event, Place, and Time details as compared to Controls (all *p* values < .01; Supplementary Table 5). The AD group, by contrast, produced significantly fewer Event details relative to Controls (*p* < .001). Between patient group comparisons did not reveal any significant differences in the types of contextual details retrieved during Free Recall (all *p* values > .1; Supplementary Table 5). Importantly, we did not find evidence of elevated external details in patients relative to Controls (all *p* values > .1; Table 2), suggesting that patients did not provide extraneous information to compensate for the paucity of internal details recalled.

**Table 2.** Total number of internal and external details (irrespective of epoch) produced from autobiographical memory recall during Free Recall and Total Recall conditions in patient and Control groups

|  | LPA | AD | Control | Magnitude of group effect<sup>‡</sup> | LPA vs. AD (*p* value) |
| --- | --- | --- | --- | --- | --- |
| *Internal details* |  |  |  |  |  |
| Free Recall | 58.6 (20.9) | 58 (25.4) | 110.4 (38.8) | $F = 14.2$; $p < .001$; $\eta_p^2 = .41$ | .78 |
| Total Recall | 159 (61.4) | 143.3 (38.3) | 221.3 (43.7) | $F = 11.2$; $p < .001$; $\eta_p^2 = .36$ | .54 |
| *External details* |  |  |  |  |  |
| Free Recall | 96.2 (39.9) | 105.3 (67.7) | 81.5 (45.0) | $F = .67$; $p = .51$; $\eta_p^2 = .03$ | .76 |
| Total Recall | 205 (72.1) | 155.2 (88.5) | 119.9 (38.4) | $F = 4$; $p = .02$; $\eta_p^2 = .16$ | **.007** |

*Note.* Free Recall score = Free Recall + General Probe condition scores. Total Recall score = Free Recall + General Probe + Specific Probe condition scores. Scores represent totals across all time periods. For all groups, mean and standard deviation reported; Magnitude of group effect measured using ANCOVAs covarying for overall language performance with *F*-statistics, exact *p*-values (unless $p < .001$ ) and effect size ( $\eta_p^2$ ) values reported; all *p*-values bolded if $p \leq .05$ ; <sup>‡</sup>For all *F*-statistics, $df_{\text{numerator}} = 2$ and $df_{\text{denominator}} = 40$ ; LPA = Logopenic Progressive Aphasia; AD = Alzheimer's disease.

**Figure 1.**
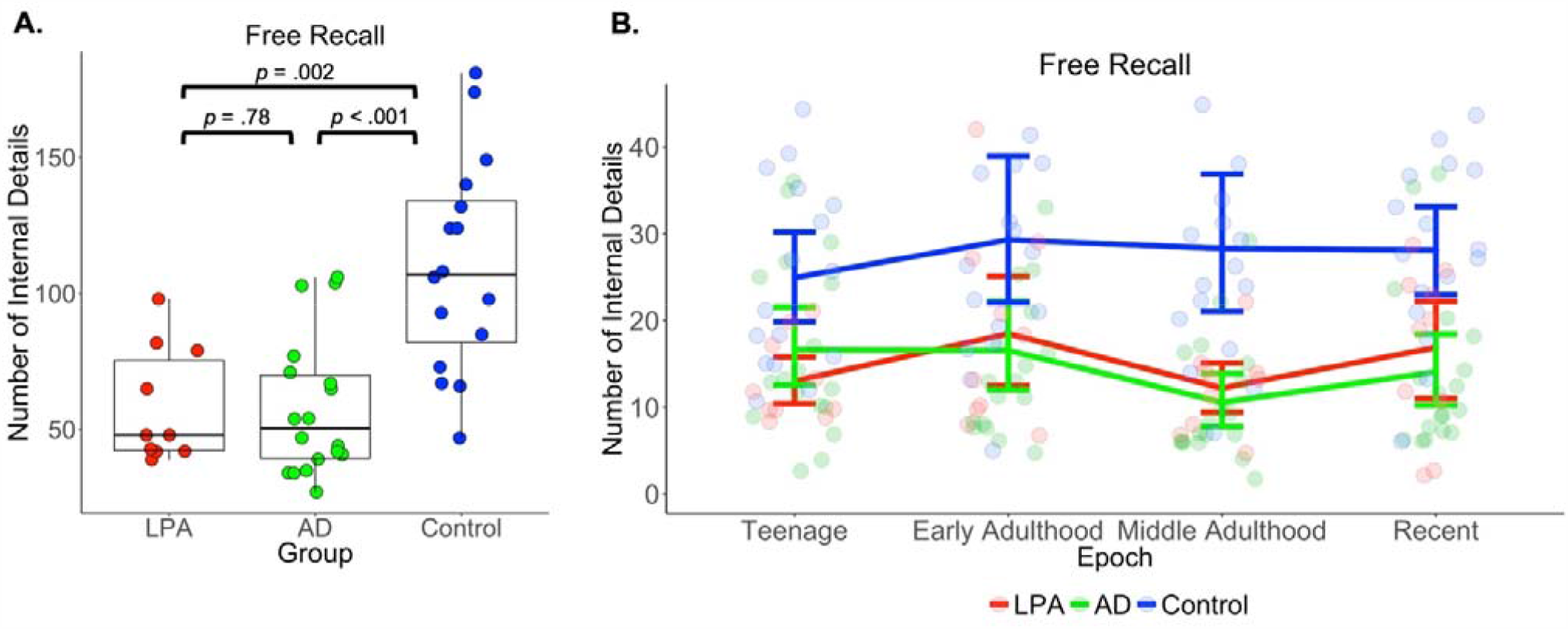
Panel A) Free Recall of internal (episodic) details, summed across time periods, on the Autobiographical Interview. Box height depicts distribution of data with lower and upper ends of the box depicting inter-quartile ranges. Bolded horizontal lines within boxes depict median score while whiskers depict the variability in distribution outside the upper and lower quartiles. Panel B) Free Recall of internal (episodic) details, broken out by time period, on the Autobiographical Interview. Error bars represent standard error of the mean. Free Recall score = Free Recall + General Probe condition scores. LPA = Logopenic Progressive Aphasia; AD = Alzheimer’s disease.

#### Free Recall across epochs

Figure 1B depicts Free Recall performance across time periods on the Autobiographical Interview for all participant groups. A repeated measures ANCOVA (controlling for overall language performance) revealed a significant main effect of Group [*F* (2, 41) = 12.7; *p* < .05; 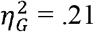], with LPA and AD recalling significantly fewer internal details relative to Controls, irrespective of time period (Figure 1B). No significant main effects were observed for Epoch [*F* (3, 123) = 1.52; *p* > .1; 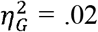], and the interaction between Group and Epoch was not significant [*F* (6, 123) = .78; *p* > .1; 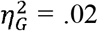]. As such, both patient groups displayed a flat retrieval profile for internal details under conditions of low retrieval support (Figure 1B).

#### Overall Total Recall

Group level performance for Total Recall of internal details (i.e., sum of Free and Probed Recall) is displayed in Table 1 and Figure 2A. A main effect of Group, controlling for overall language dysfunction, was noted [*F* (2, 40) = 11.2, *p* < .001, 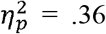] driven by poor performance in LPA and AD relative to Controls (*p* < .05), with no differences between the patient groups (*p* > .1) (Figure 2A, Table 2). A significant group effect was also found for the proportion of internal details generated within the autobiographical narrative, covarying for language performance [*F* (2, 40) = 15.3; *p* < .001; 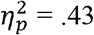], with both patient groups scoring significantly lower than Controls (both *p* values < .001; Supplementary Table 4) and no differences between the patient groups (*p* = .17). The types of internal details generated following probing further differed between the groups (Supplementary Table 5). While the LPA group scored in line with Controls for Event, Perceptual, and Thought/Emotion details (all *p* values > .07), they generated fewer Place and Time details despite the provision of structured probes (both *p* values < .05). In contrast, the AD group produced significantly fewer Event, Place, Perceptual, and Thought/Emotion details relative to Controls (all *p* values < .05), and no significant differences were evident between the patient groups across contextual detail subcategories (all *p* values > .06; Supplementary Table 5). Finally, a significant group effect was found for external details [*F* = 4; *p* = .02; 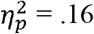] reflecting the increased provision of off-target information in LPA relative to Controls following structured probing (*p* = .007). No other significant differences were evident (all *p* values > .1) (Table 2).

**Figure 2.**
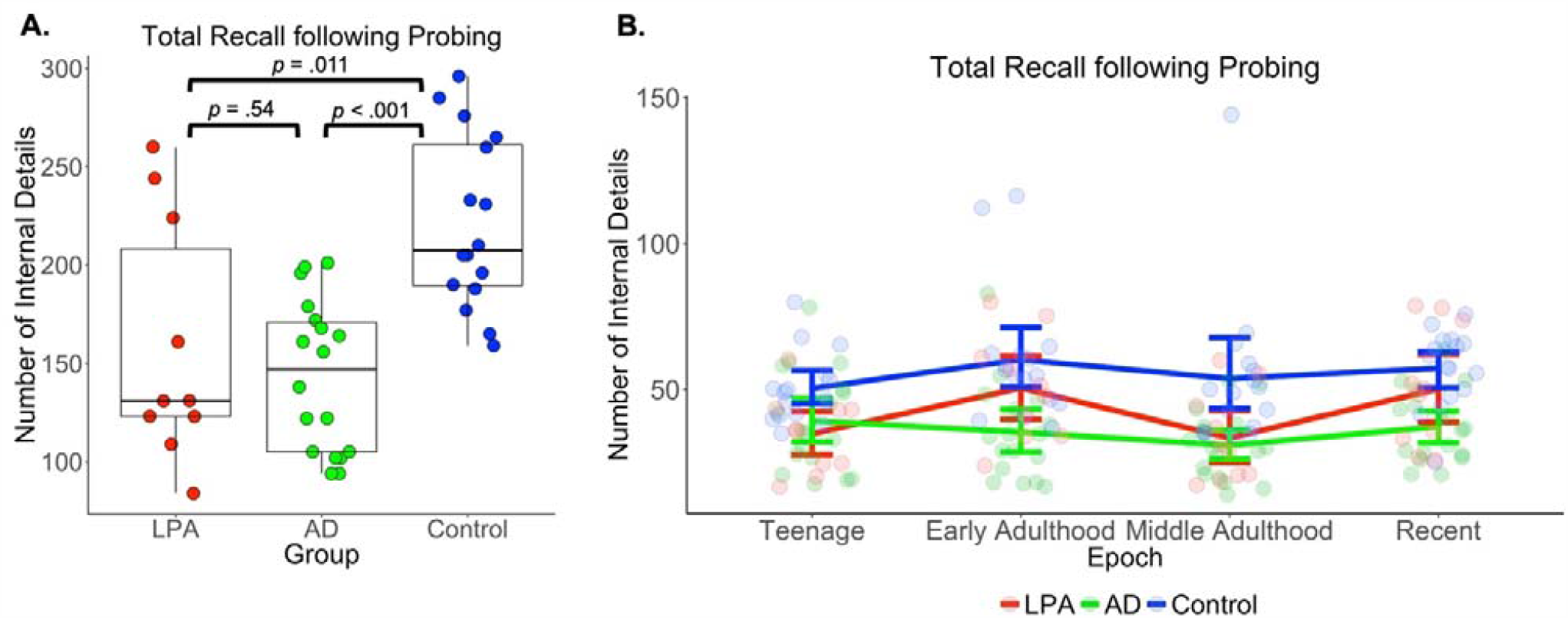
Panel A) Total Recall of internal (episodic) details, summed across time periods, on the Autobiographical Interview. Box height depicts distribution of data with lower and upper ends of the box depicting inter-quartile ranges. Bolded horizontal lines within boxes depict median score while whiskers depict the variability in distribution outside the upper and lower quartiles. Panel B) Total Recall of internal (episodic) details, broken out by time period, on the Autobiographical Interview. Error bars represent standard error of the mean. Total Recall score = Free Recall + General Probe + Specific Probe condition scores. LPA = Logopenic Progressive Aphasia; AD = Alzheimer’s disease.

#### Total Recall across epochs

A repeated measures ANCOVA, covarying for overall language performance, was run to explore Total Recall of internal details following structured probing across the four time periods of the Autobiographical Interview. A significant main effect of Group [*F* (2, 41) = 11.7; *p* < .05; 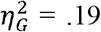] was found, reflecting overall diminished internal detail retrieval in LPA and AD, irrespective of epoch, compared to Controls (Figure 2B). A significant main effect of Epoch was also observed [*F* (3, 123) = 4.15; *p* < .05; 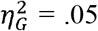] whereby all groups demonstrated significantly higher levels of internal details in Early Adulthood (Figure 2B) relative to all other epochs. The interaction between Group and Epoch, however, was not significant [*F* (6, 123) = 1.6; *p* > .1; 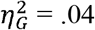).

#### Autobiographical memory deficits in LPA are not related to disease severity or global cognitive status

Pearson’s correlations were run to explore associations between autobiographical memory performance and disease severity metrics in both patient groups. In both LPA and AD groups, autobiographical memory performance was not found to correlate significantly with clinician-indexed disease severity (CDR-FTLD SoB) (all *p* values > .1; Supplementary Table 6).

To further ensure that autobiographical memory impairment was not simply a product of more severe cognitive dysfunction in LPA, we used a median split based on the ACE-R Total score (median score = 75.0) to create two LPA subgroups (High ACE-R, *N* = 5; Low ACE-R, *N* = 5). There was no evidence for any significant group differences between high and low ACE-R Total subgroups for Free Recall (*t* = -.89; *p* = .40) or Total Recall (*t* = -.49; *p* = .63) performance. As such, autobiographical memory retrieval disturbances in LPA cannot be explained by more advanced disease staging or by the overall severity of cognitive impairment.

#### Associations between autobiographical memory and general cognitive performance

In the LPA group, Pearson’s correlations revealed no significant associations between Free or Total Recall of internal details and targeted measures of neuropsychological performance (including tests of language abilities) (all *p* values > .1; Supplementary Table 6). In AD, Free Recall of internal details was associated with auditory attention (Digit Span Forward, *r* = .68; *p* = .04), whereas Total Recall was associated with a global measure of episodic memory function (ACE-R Memory subscale, *r* = .68; *p* = .05; Supplementary Table 5).

### VBM analyses

#### Group differences in grey matter intensity

Profiles of grey matter intensity decrease in the LPA and AD groups, relative to Controls, are detailed in Supplementary Results, Supplementary Table 7, and Supplementary Figure 1. Briefly, LPA patients displayed canonical grey matter intensity reduction centred on the left IPL and peri-sylvian temporal cortices, extending medially into the posterior portions of the left hippocampus, and including bilateral medial prefrontal, left insular, left postcentral gyrus and occipitopolar regions. AD patients, by contrast, displayed a bilateral pattern of medial temporal and hippocampal (across the longitudinal axis), lateral and medial parietal, and lateral temporal cortical atrophy, extending to the bilateral prefrontal and insular cortices. These patterns of atrophy are in keeping with previous independent reports in the literature for LPA (Gorno-Tempini *et al*., 2008; Teichmann *et al*., 2013; Rogalski *et al*., 2014) and AD (Karas *et al*., 2004; Moller *et al*., 2013).

#### Grey matter correlates of autobiographical retrieval

##### Free recall in LPA

In LPA, Free Recall difficulties were associated with grey matter intensity decrease predominantly in the left angular gyrus encroaching dorsally to the left superior parietal cortex and left precuneus. A right parieto-occipital cluster was also found, encompassing the right lateral occipital and supracalcarine cortices and the right precuneus. Finally, a third cluster involving the right inferior frontal gyrus and the precentral gyrus was observed (Figure 3 and Table 3).

**Table 3.**
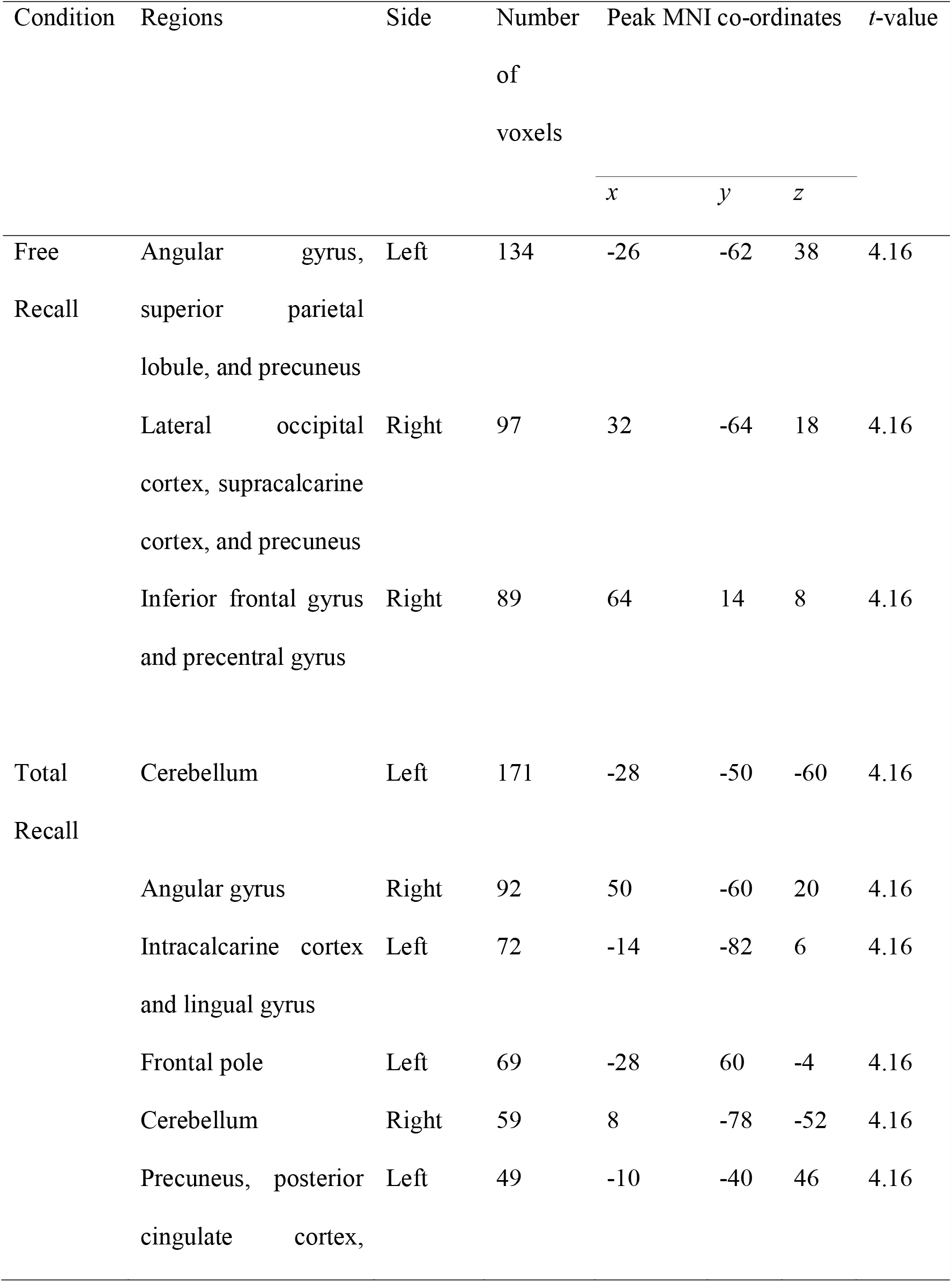
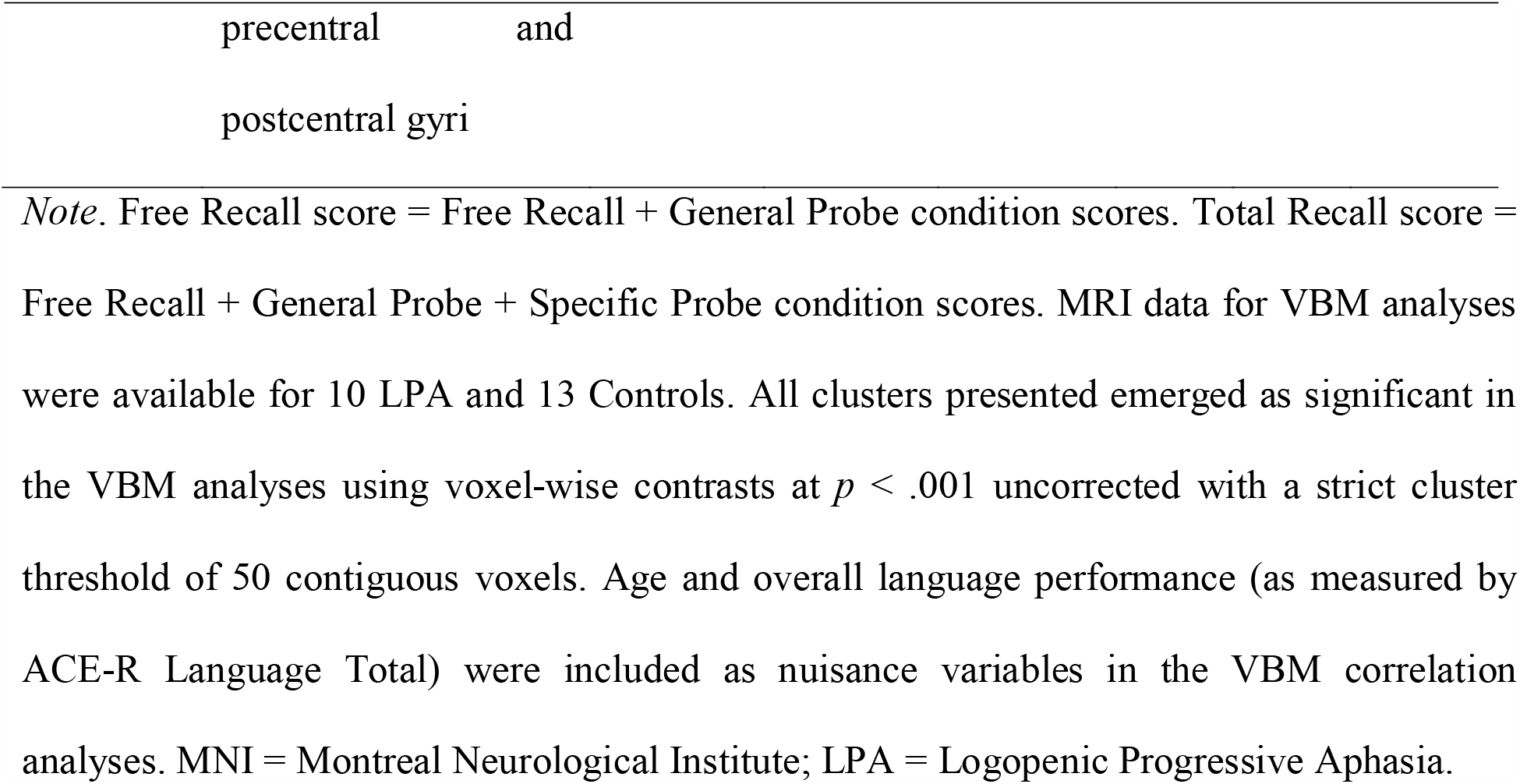
Voxel-based morphometry results showing regions of significant grey matter intensity decrease that correlate with total recall of internal details from autobiographical memory on Free Recall and Total Recall conditions in LPA and Control groups.

**Figure 3.**
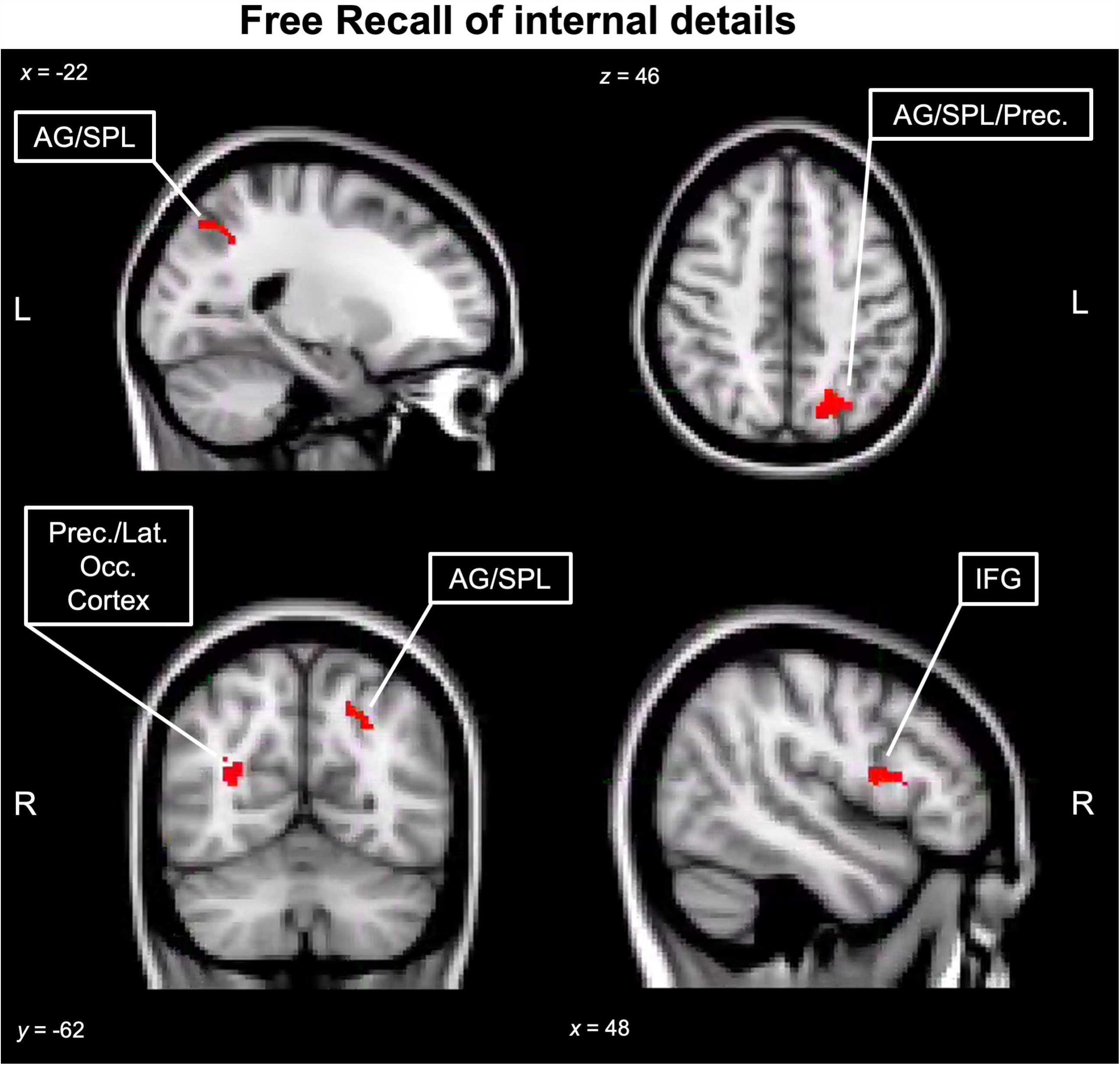
Regions of significant grey matter intensity decrease that correlate with reduced Free Recall of internal details on the Autobiographical Interview in LPA and Controls. Free Recall score = Free Recall + General Probe condition scores. Coloured voxels indicate regions that emerged as significant in the voxel-based morphometry analyses using voxel-wise contrasts at *p* < .001 uncorrected for multiple comparisons with a strict cluster threshold of 50 contiguous voxels. All clusters reported at *t* = 4.16. Age and overall language performance (measured using the ACE-R Language Total score) were included as nuisance variables. Clusters are overlaid on the Montreal Neurological Institute (MNI) standard brain with *x, y*, and *z* coordinates reported in MNI standard space. L = Left, R = Right, LPA= Logopenic Progressive Aphasia, AG = Angular Gyrus, SPL = Superior Parietal Lobule, Prec. = Precuneus, Lat. Occ. Cortex = Lateral Occipital Cortex, IFG = Inferior Frontal Gyrus.

#### Total Recall in LPA

Total internal details retrieved following structured probing in LPA was associated with grey matter intensity in the right angular gyrus, left posterior parietal cortices including the precuneus, posterior cingulate cortex, and pre/postcentral gyri, as well as left occipital regions including the intracalcarine and lingual gyri. Smaller clusters in the bilateral cerebellar cortices and the left frontal pole were also observed (Figure 4 and Table 3).

**Figure 4.**
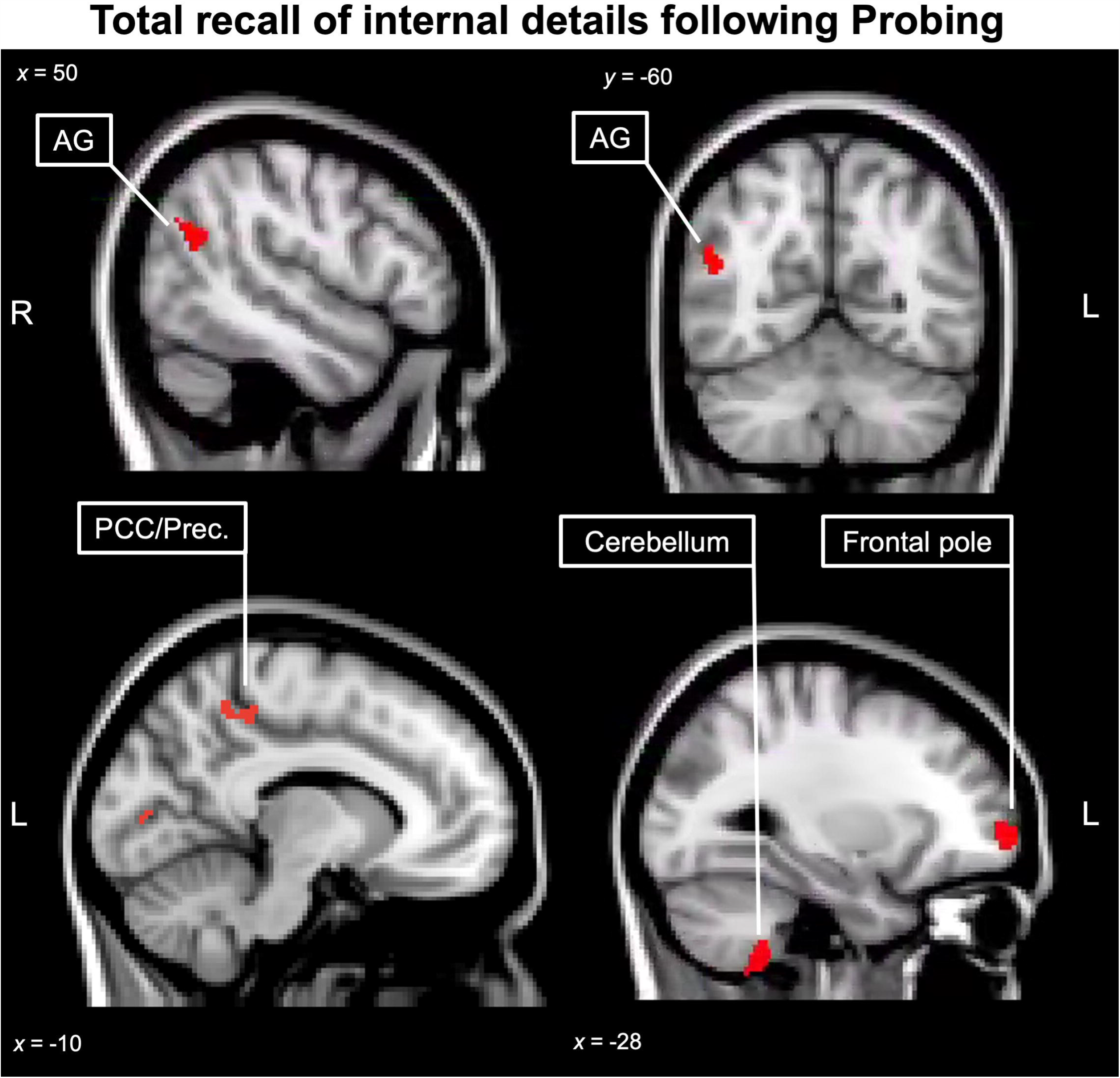
Regions of significant grey matter intensity reduction that correlate with Total Recall of internal details on the Autobiographical Interview in LPA and Controls. Total Recall score = Free Recall + General Probe + Specific Probe condition scores. Coloured voxels indicate regions that emerged as significant in the voxel-based morphometry analyses using voxel-wise contrasts at *p* < .001 uncorrected for multiple comparisons with a strict cluster threshold of 50 contiguous voxels. All clusters reported at *t* = 4.16. Age and overall language performance (measured using the ACE-R Language Total score) were included as nuisance variables. Clusters are overlaid on the Montreal Neurological Institute (MNI) standard brain with *x* and *y* coordinates reported in MNI standard space. L = Left, R = Right, LPA= Logopenic Progressive Aphasia, AG = Angular Gyrus, Prec. = Precuneus, PCC = Posterior Cingulate Cortex.

#### Free Recall in AD

In AD, reduced Free Recall was associated with grey matter intensity decrease in a distributed set of regions including the right posterior hippocampus, bilateral posterior parietal (IPL, precuneus, and superior parietal lobes) and inferior frontal cortices. The left insula, orbitofrontal cortex, and temporal pole, as well as right thalamus, caudate nucleus, and cerebellum also emerged as significant in the analysis (Supplementary Figure 2 and Supplementary Table 8).

#### Total Recall in AD

Total Recall of internal details in AD was associated with a distributed network of medial temporal, parietal and frontal regions including bilateral posterior hippocampi, right middle temporal and IPL regions, left insular, and bilateral frontopolar and ventromedial prefrontal cortices (Supplementary Figure 3 and Supplementary Table 8).

## DISCUSSION

Although widely characterised as a disorder of language, accumulating evidence indicates marked episodic memory difficulties on standardized neuropsychological tasks in LPA (Eikelboom *et al*., 2018). Here, we demonstrate for the first time that episodic memory impairments in LPA extend to the domain of autobiographical memory. In a well-characterized LPA group, all of whom presented with language output problems as their primary and predominant symptom, we reveal a global disruption in autobiographical retrieval in LPA, spanning recent and remote time periods. This observation of a flat gradient of retrieval across four distinct life epochs, of the same magnitude as that observed in disease-matched cases of AD, suggests that autobiographical memory impairments in LPA do not simply reflect post-disease onset encoding difficulties. By controlling for overall level of language dysfunction in our analyses, we further show that autobiographical retrieval difficulties cannot be attributed to lexical retrieval and overall language dysfunction characteristic of these patients. Moreover, while LPA patients benefited from the provision of structured probing, this was not sufficient to bring their performance in line with that of Controls. We also found no significant associations between autobiographical performance in either condition with clinician-indexed disease severity in the LPA group, strengthening our proposal that episodic memory impairments are a core feature of LPA, regardless of disease staging. Finally, we revealed that autobiographical memory deficits are present in LPA patients, irrespective of their overall level of cognitive decline. Taken together, our findings reveal the presence of a genuine autobiographical amnesia in LPA that cannot be explained by disease severity, overall cognitive status, language dysfunction, post-disease onset encoding difficulties, or strategic retrieval deficits.

Whole-brain VBM analyses enabled us to map associations between changes in grey matter intensity and Free Recall and Total Recall of internal (episodic) details. After covarying for overall language performance, impoverished Free Recall in LPA was associated with grey matter intensity decrease predominantly in left posterior parietal regions, most notably the left angular gyrus, left superior parietal lobule, and the bilateral precuneus. The focal point of maximal grey matter degeneration in LPA resides in the left temporoparietal cortices, and is a major contributor to lexical retrieval and verbal working memory dysfunction in the syndrome (Leyton *et al*., 2012). Accordingly, the emergence of episodic memory difficulties in LPA has typically been attributed to primary lexical retrieval difficulties, mediated by lateral temporal degeneration (Win *et al*., 2017). In contrast, our previous work indicate that the degeneration of posterior parietal cortices in LPA produces a stark episodic amnesia (Ramanan *et al*., 2020). Here we extend these findings by demonstrating the domain-general role for the IPL in retrieving personally-relevant episodic details from the remote past (Ramanan and Bellana, 2019). These results resonate with functional neuroimaging studies that consistently implicate the IPL and precuneus as core nodes of a distributed episodic retrieval brain network, further specialized in processing contextual details from autobiographical memory (reviewed by Ramanan *et al*., 2018; Ritchey and Cooper, 2020). Importantly, our VBM analyses further indicated involvement of the right inferior frontal gyrus in Free Recall disturbances in LPA, in line with a large body of work showing inferior frontal and prefrontal involvement in strategic search, memory control, and inhibition of irrelevant information during autobiographical recall (Simons and Spiers, 2003; Aron *et al*., 2014; Diamond and Levine, 2018).

Reduced Total Recall performance in LPA was found to correlate with grey matter intensity decrease in left medial parietal regions (precuneus and posterior cingulate cortex), angular gyrus in the right IPL, as well as left frontopolar and bilateral cerebellar regions. Although not typically affected early in the LPA disease trajectory, these regions border anterior perisylvian, ventral temporal, and medial parietal cortices, which, in themselves, are early sites of pathology in the syndrome (Ossenkoppele *et al*., 2015). Moreover, these regions are known to support a variety of cognitive processes that aid the successful retrieval of autobiographical memories. For example, the posterior cingulate cortex plays an established role in the reinstatement of self-referential information from memory (Northoff and Bermpohl, 2004; Bird *et al*., 2015), while the right IPL is a key hub of the bottom-up attentional network (Corbetta *et al*., 2008) capturing relevant information within autobiographical memory in response to structured probes (Cabeza, 2008; Cabeza *et al*., 2008). Our observation of an increase in non-episodic (external) details in LPA following structured probing may reflect diminished episodic control over memory producing a general inefficiency in autobiographical retrieval (see also Esopenko and Levine, 2017; Carmichael *et al*., 2019). Our finding of cerebellar involvement is in keeping with recent work outlining the role of the cerebellum in sequencing information for subsequent autobiographical retrieval (Addis *et al*., 2016) and a growing literature implicating the cerebellum across a wide range of cognitive disturbances in neurodegenerative disorders (Chen *et al*., 2018; Jacobs *et al*., 2018). As such, the impaired capacity to retrieve contextual information following structured probing in LPA likely reflects the breakdown of multiple interacting processes including searching, monitoring, and retrieval of appropriate information, as well as cognitive control processes.

While we focus here on the relationship between posterior parietal lobe dysfunction and autobiographical memory impairments, it is important to note that the precise functional contributions of subregions within the posterior parietal cortices to episodic memory remain to be clarified. In this regard, it would be interesting to compare memory profiles in LPA with Corticobasal Syndrome and Posterior Cortical Atrophy – two syndromes that present with early motor and visual complaints, respectively, amidst primary parietal cortex degeneration. Notably, recent work has revealed the presence of marked episodic memory difficulties attributable, in part, to posterior parietal dysfunction in both Corticobasal Syndrome (Ramanan *et al*., 2019b) and Posterior Cortical Atrophy (Ahmed *et al*., 2018). Teasing apart different aspects of memory dysfunction in these populations may provide important clues regarding the functional role of discrete regions of the posterior neocortex.

A further point to consider is that we did not find significant associations between the hippocampus and Free or Total Recall from autobiographical memory. This stands in contrast with our previous study in which the left hippocampus emerged as associated with verbal episodic memory performance in LPA (Ramanan *et al*., 2020). One potential explanation is that, unlike our previous study, only a relatively small portion of the left posterior hippocampus was affected in the LPA cohort included here. In contrast, widespread atrophy across the longitudinal axis of the hippocampus and medial temporal lobes more broadly is not typically noted until later stages of LPA (Phillips *et al*., 2018). Furthermore, the neural correlates supporting episodic retrieval and autobiographical recall vary, with autobiographical retrieval tending to greatly engage parietal regions that aid in the processing of detailed contextual information (Chen *et al*., 2017; Ramanan, 2017). While our findings suggest that predominantly posterior parietal network damage significantly disrupts autobiographical retrieval in LPA, longitudinal studies will be particularly important to determine how the encroachment of atrophy into medial temporal lobe regions impacts discrete aspects of the recollective endeavour.

A number of methodological considerations warrant discussion. Due to the rarity of the LPA syndrome and the time-consuming, but highly detailed, nature of the Autobiographical Interview (∼2 hours administration time per person), we limited our LPA patient pool to ten well-characterised cases with accompanying comprehensive neuropsychological and neuroimaging data. This provides us with a richly detailed dataset giving rise to the unique findings reported here. Our LPA cohort was diagnosed strictly in line with current diagnostic criteria (Gorno-Tempini *et al*., 2011), however, the majority of our sample have not yet come to autopsy. This prevents us from exploring potential associations between memory performance and underlying pathology in LPA – an avenue that future studies employing larger LPA cohorts should explore. Next, we report our grey matter correlational analyses at a conservative uncorrected threshold of *p* < .001, however, given our use of strict cluster thresholds and the strong convergence between our findings and a large number of independent studies, we are confident that our results do not represent false positives. Finally, we limited our analyses to grey matter morphometric changes, however, future studies should also include diffusion-weighted imaging to determine how alterations in structural connectivity between parieto-hippocampal regions relate to profiles of memory impairment in LPA.

Our findings hold a number of clinical implications for the accurate diagnosis and characterisation of LPA. Given its conceptualisation as a primary aphasic disorder, the presence of remote and recent memory difficulties in LPA may unwittingly bias clinicians to consider a diagnosis of typical AD with language features. This risk is compounded in LPA patients that present with marked amnesia in the context of amyloid-positive biomarkers. Recent evidence suggests that linguistic markers such as lexical retrieval difficulties, thought to be unique to LPA, may in fact be present across primary progressive aphasia syndromes (Giannini *et al*., 2017), limiting the clinical utility of language performance as the sole neuropsychological criterion in differentiating the primary progressive aphasias. On the other hand, early episodic memory impairment amongst primary progressive aphasias is thought to be unique to LPA (Ramanan *et al*., 2016; Eikelboom *et al*., 2018), indicating the need to consider the concurrent presentation of language and memory disturbances when diagnosing this syndrome. Future research should examine the clinical utility of episodic memory performance in differentiating LPA patients with amyloid-positive versus amyloid-negative profiles with the overall aim to improve clinicopathological prediction, thereby directly aiding clinical trial development.

In summary, this is the first study, to our knowledge, to demonstrate stark impairments of recent and remote autobiographical retrieval in LPA, that cannot be attributed to disease severity, overall cognitive status, language dysfunction, post-disease encoding difficulties, or strategic retrieval deficits. Our findings reveal a new dimension of cognitive impairment not previously associated with the clinical presentation of LPA and suggest a need to rethink our approach towards the conceptualisation and diagnosis of this syndrome.

## ACKNOWLEDGEMENTS

The authors are grateful to the patients and families for their continued support of our research. The authors wish to acknowledge the Sydney Informatics Hub funded by the University of Sydney for providing access to High Performance Computing (HPC) facilities.

## SUPPLEMENTARY MATERIAL

1 Supplementary file with Supplementary Methods and Results, 8 Supplementary Tables, and 3 Supplementary Figures.

